# Disentangling Blood-Based Markers of Multiple Sclerosis Through Machine Learning: An Evaluation Study

**DOI:** 10.1101/2025.04.02.25325148

**Authors:** Robin Vlieger, Mst Mousumi Rizia, Abolfazl Amjadipour, Nicolas Cherbuin, Anne Brüstle, Hanna Suominen

## Abstract

In the search for markers to aid early diagnosis, sustainable monitoring, and accurate prognosis of Multiple Sclerosis (MS), researchers have turned to blood-based markers. These provide rich information on a person’s health while being easier to acquire than magnetic resonance images. To analyse blood data, researchers have used machine learning (ML) to support evaluation at scale, but because many different analytics pipelines exist, it is unclear how different ML methods compare and influence experimental outcomes. Therefore, this ML evaluation study compared in different configurations the performance of five ML algorithms, two methods to select their features, and approaches to evaluate them. The aim was to first assess how the ML methods influenced classifying people with MS and controls, and then disentangle the blood-based markers selected for the best performing classifiers. The results indicated that Logistic Regression with Random Forests for feature selection and 10-fold cross-validation produced the best results, that feature selection depended on the feature selection methods, and that data splits for training, validation, and testing were heterogeneous. This suggests experimental setups influence both the classification performance and disentangled markers, meaning that evaluation rigor matters when using ML to support discovery processes and knowledge creation in medical research.

## 1. Introduction

Multiple Sclerosis (MS) is a neurodegenerative condition for which diagnosis [1], monitoring, and prognosis can be difficult. Diagnosis, based on Magnetic Resonance Imaging (MRI), requires the presence of brain lesions that appear at different times and in different parts of the brain [1]. This requirement leads to delays in diagnosis and commencement of treatment. Development of the disease is highly variable between individuals, hindering accurate prognosis. Additionally, MRI scanners are expensive, leading to them generally only being available in densely populated areas. For People with MS (PwMS) outside of such areas, this results in challenges in diagnosis and MS monitoring from additional travel, which is undesirable as many struggle with fatigue. This has led to research into more affordable, more easily accessible, and accurate markers of MS.

As dysregulation of the immune system in MS can be observed through blood samples, researchers have investigated them as potential markers of MS [2]. They provide a wealth of information on a person’s well-being, with the combination of different measurements, such as cell counts and ratios, and interactions being of special interest. Because of their high-dimensional nature, investigators have applied Machine Learning (ML) to the data due to its capability to find relationships between variables (called features in ML) and identify combinations that lead to accurate outcomes in, for example, classifying those with versus those without MS or extrapolating MS severity, respectively [3,4]. The ML methods employed vary from conventional algorithms such as Random Forests (RFs)[5] and Support Vector Machines (SVMs)[6] to Deep Learning (DL) methods such as Artificial Neural Networks (ANNs)[7]. Unfortunately, the myriad of ML methods in different studies makes comparing and interpreting their results problematic.

To clarify how methodological choices influence experimental outcomes, a comparative evaluation study was undertaken. Namely, the performance of five ML algorithms, two methods to select their features, and three Cross-Validation (CV) approaches compared in different configurations. Female PwMS and female controls were classified based on blood-based markers, using 10-fold (10F), leave-one-out (LOO), or leave-two-out (LTO) CV, either a RF Classifier (RFC) or t-test for feature selection, and a Logistic Regression Classifier (LRC)[8], SVM, Gradient Boosting Classifier (GBC)[9], Feed Forward Neural Network (FFN), or Transformer architecture (TRF) as the algorithm. Combining LRC, RFC, and 10F CV produced the best performing classifier. The results also indicated that disentangled markers of MS depend on both combined ML methods and data for their training, validation, and testing.

## 2 Methods

Data were collected at the Australian National University (ANU) as part of an ongoing longitudinal cohort study by the ‘Our Health in Our Hands’ (OHIOH) strategic initiative [10]. This study was approved by the ACT Health (2019.ETH.00081) and ANU Human Research Ethic Committees (2020/047), currently chaired by Prof Paul Gatenby Prof and Michelle Banfield, respectively, and informed written consent was obtained from all participants. Bloods were drawn when participants entered the study and every subsequent year. From those data, 21 female PwMS and 21 controls were age-matched (average age 50.1 years ± 12.2) in the study as MS is more prevalent in females and to avoid any sex-based confounds.

Five different algorithms including LRCs, SVMs, GBCs, FFNs, and TRFs were used. The LRC was selected for its simplicity, and the SVM due to its popularity in medical ML classification tasks [6]. The GBC was chosen for its excellent performance in classification competitions [11] and its capacity to handle non-normally distributed data. 1-layer FFNs were used after preliminary tests showed better results than FFNNs with more layers, as a relatively small DL architecture, and TRFs [12] were included due to their popularity in DL research.

A nested CV approach [13] was deployed, using one of 10F, LTO, or LOO CV protocols, for the inner and outer loop. For example, in the 10F experiments, the data were divided into 10 equal sets. 90% of the data was used in the inner loop for training and validation, and the final model was tested on the last 10%. This was repeated until each set had been the test set. To avoid data leakage, normalisation, and feature selection were implemented as part of the training process, using Scikit-Learn’s PowerTransformer implementation. Either an RFC or t-test (i.e., feature ranking by Gini-coefficients [14] or p-values, respectively) was applied to selecting features. The top 7 features were used to build the model. For each algorithm, all three CV approaches were assessed, while feature selection methods were assessed for LRCs and SVMs. For GBCs, being a tree method, no feature selection was required, nor for the two DL algorithms. See the online supplement for the list of features, their abbreviations, and their ranks per experiment, as well as for hyperparameters that were optimised and their value ranges.

Accuracy was used as the primary measure of overall performance, and sensitivity, specificity, precision, and F1-score were obtained to further evaluate performance, and Analyses of Variance (ANOVAs) were used to test for statistically significant differences between models, using a Bonferroni-corrected significance level of 0.01. To determine which features were determined by the models to distinguish PwMS from controls most accurately, Gini-coefficients and p-values were extracted from the models for all features.

## 3. Results

Differences in evaluation results were explained by both the ML/DL algorithm and CV approach (Table 1). LRC experiments gave the highest scores on the evaluation metrics when using 10F CV. Evaluation scores were also highest for the 10F CV experiments when using TRF, GBC, and SVM algorithms, regardless of their feature selection method. In the FFN experiments, the LTO CV produced higher scores. Specificity was higher than sensitivity in almost every experiment. Based on ANOVAs, specificity (*F*(4, 515) = 3.81, p < 0.01), precision (*F*(4, 515) = 6.11, p < 0.01), and F1(*F*(4, 515) = 4.60, p < 0.01) produced statistically significant differences between groups.

**Table 1.** Evaluation metrics (Accuracy (Acc), sensitivity, specificity, precision, and F1) in percentages for the 5 best performing experiments when comparing classification algorithms, Feature selection methods (Feat.), and CV approaches. Best scores are in **bold**.

| Algorithm | Feat. | CV | Train Acc | Test Acc | Sensitivity | Specificity | Precision | F1 |
| --- | --- | --- | --- | --- | --- | --- | --- | --- |
| GBC | - | 10F | 69.87 | 63.55 | **62.94** | 65.30 | 66.46 | **63.01** |
| FFN | - | LTO | 66.80 | 61.37 | 52.74 | 66.59 | 55.36 | 50.65 |
| TRF | - | 10F | 58.64 | 57.85 | 58.83 | 58.90 | 59.09 | 56.18 |
| LRC | RFC | 10F | 63.96 | **66.36** | 62.89 | **73.14** | **72.40** | 63.00 |
| SVM | RFC | 10F | 63.29 | 58.44 | 59.72 | 58.00 | 56.19 | 55.63 |

In the LRC experiment with RFC and 10F CV large differences in per-fold test scores were observed. One test fold only produced an accuracy of 19.73%, but two test folds produced accuracies of 87.50%. The heterogeneity was also seen in the scores for sensitivity and specificity, with one test fold producing 100.00% sensitivity scores and another 97.64%, while three test folds produced perfect specificity scores. High sensitivity did not mean a drop in specificity or vice versa. Four folds produced F1-scores higher than 72.00%. Similar disparities also occurred for other algorithms.

Comparing feature selection methods, t-tests produced better results for the LRC with LTO or LOO CV combinations, but combining RFCs worked better for LRCs/SVMs and 10F CV. Employing LRCs for classification with either feature selection method scored better in evaluation than using SVMs. The largest accuracy difference of 7.93% between LRCs and SVMs was found with RFCs and 10F CV while the smallest, being 0.73%, occurred for RFCs and LOO CV. Most important blood-based features varied in method combinations. For the GBC algorithm, they were EOS%, *EOS*, BAS%, RDW, BAS, MPV, and MCHC. For RFCs for feature selection, the list changed to LYM%, NEU%, LYM, MCH, MCV, WBC, and MPV, while t-tests selected LYM%, NEU%, LYM, MCH, MCV, EOS%, and *EOS*.

## 4. Discussion

This study clarified effects of methodological choices on outcomes in a task classifying females with MS and controls based on blood values. Outcomes differed based on choice of algorithm, CV method, and feature selection method. Statistically significant differences were observed on the evaluation metrics specificity, precision and F1-score, and selected features differed between feature selection methods. Heterogeneous values for evaluation scores from individual folds indicate differences exist between participants.

Our findings are limited by cohort choice, as we used 21 age-matched pairs of female participants with an average age of 50 years. The results cannot be extrapolated to a population consisting of people of both sexes or different ages. Additionally, our choices limited the number of participants, which may have influenced results, as a larger cohort might lead to less heterogeneous evaluation scores. The DL methods did not perform better than conventional algorithms, which can be ascribed to data scarcity.

Variance in evaluation scores for individual test sets suggest an influence of data division. Further research could focus on the instance classification, meaning the classification of the individual participants, and identify what makes that a person is classified correctly or incorrectly, not only based on the features available in the dataset, but also the clinical and demographic information if this is available.

Our study did not intend to produce a state-of-the-art ML method for diagnosis of MS, and other studies, often using other data modalities, have achieved higher evaluation scores. While true, comparison of results is difficult: the review by Aslam et al. [3], reports on a wide variety of methodological choices and setups. This study fits in with previous studies by elucidating the effects methodological choices have on outcomes.

In a broader context, our study adds to a larger corpus of research into evaluation of ML methods [15,16,17]. Such studies have identified ML methods that produced the best results in specific context, which may not work in another context. As results seem to depend on experimental setup, an ensemble approach, meaning the combination of algorithms and feature selection methods, might also lead to improved performance.

Based on the findings in our study and previous studies [13], a nested CV approach should be recommended. Which CV approach is used within the nested CV approach can be a methodological choice, but does need clarification, as it can influence results. Additionally, researchers should be aware of the effects feature selection methods might have on the identification of potential markers. If marker discovery is the goal, an ensemble method might be preferred to avoid bias of just one method recommending a feature.

## 5. Conclusion

This study makes clear that methodological choices influence outcomes when classifying PwMS and controls using blood values. For blood values to become a viable and cost-effective option as a marker for diagnosis and prognosis in MS, also alleviating the burden on PwMS by improving accessibility and helping with fatigue management, it is necessary for future research to argue clearly why methods were chosen, so it can be identified what leads to differences, and an ensemble approach might improve overall performance if marker discovery is the goal.

## Data Availability

All data produced in the present study are available upon reasonable request to the authors.

https://github.com/RobinVliegerAcademic/MedInfo2025_Vlieger_et_al

## Footnotes

This research was funded by the ANU OHIOH initiative, which aims to transform healthcare by developing new personalised health technologies and solutions in collaboration with patients, clinicians, and healthcare providers.

